# Efficacy of a multiple-component and multifactorial personalized fall prevention program in a mixed population of community-dwelling older adults with stroke, Parkinson’s disease, or frailty compared to usual care: The PRE.C.I.S.A. Randomized Controlled Trial

**DOI:** 10.1101/2022.03.26.22272987

**Authors:** Fabio La Porta, Giada Lullini, Serena Caselli, Franco Valzania, Chiara Mussi, Claudio Tedeschi, Giulio Pioli, Massimo Bondavalli, Marco Bertolotti, Federico Banchelli, Roberto D’Amico, Roberto Vicini, Silvia Puglisi, Pierina Viviana Clerici, Lorenzo Chiari, the PRECISA Group

**Author notes:** Correspondence: Corresponding Author: Giada Lullini.

## Abstract

**Background:** Fall risk in the elderly is a major public health issue due to the injury-related consequences and the risk of associated long-term disability. However, the delivery of preventive interventions in usual clinical practice still represents a challenge.

**Aim:** To evaluate the efficacy of a multiple-component combined with a multifactorial personalized intervention in reducing fall rates in a mixed population of community-dwelling elderly, compared to usual care.

**Design:** Randomized Controlled Trial (NCT03592420, clinicaltrials.gov). Setting: Outpatients in two Italian centers.

**Population:** 403 community-dwelling elderly at moderate-to-high fall risk, including subjects with Parkinson’s Disease and stroke.

**Methods:** After the randomization, the described interventions were administered to the intervention group (n=203). The control group (n=200) received usual care and recommendations to minimize fall risk factors. Each participant received a fall diary and was followed by 12 monthly phone calls. The primary endpoint was the number of falls in each group over 12 months. The secondary endpoints were other fall-related indicators recorded at 12 months. Participants’ functioning was assessed at baseline (T1) and 3-month (T3).

**Results:** 690 falls were reported at 12 months, 48.8% in the intervention and 51.2% in the control group, with 1.66 (± 3.5) and 1.77 (± 3.2) mean falls per subject, respectively. Subjects with ≥ 1 fall were 236 (58.6%) and with ≥2 falls 148 (36.7%). No statistically significant differences were observed between groups regarding the number of falls, the falling probability, and the time to the first fall. According to the subgroup analysis, no significant differences were reported. A statistically significant difference was found for the Mini-BESTest (p=0.004) and the Fullerton Advanced Balance Scale (p=0.006) for the intervention group, with a small effect size (Cohen’s d 0.26 and 0.32, respectively), at T1 and T3 evaluations.

**Conclusions:** The intervention was ineffective in reducing the number of falls, the falling probability, and the time to the first fall at 12 months in a mixed population of community-dwelling elderly. A significant improvement for two balance indicators was recorded in the intervention group. Future studies are needed to explore different effects of the proposed interventions to reduce falls and consequences.

## 1 Introduction

Fall risk in the elderly is a major public health issue due to the immediate injury-related consequences and the risk of associated long-term disability (1). One out of three older people over 65 years is estimated to fall each year, and this rate increases to 50% in the elderly over 80 years old (2). Furthermore, around 15% of older adults are multiple fallers, experiencing more than one fall each year, thus increasing morbidity and mortality (1). In 2 to 10% of cases, falls can lead to hip fractures related to functional decline, death, and increased hospitalization costs, even though falls alone limit, per se, social participation and may increase the risk of institutionalization (3). Moreover, the costs for the acute management of the 85,762 hospitalizations for hip fractures that occurred in Italy in 2005 were estimated to be around 467 million Euros, with rehabilitation costs reaching 532 million Euros in the same year (4). In Regione Emilia-Romagna (Italy), Berti and colleagues (5) reported 5904 yearly hip fractures in 2017. Referring to a conceptual framework for a hip fracture integrated episode of care, defined as Continuum-Care Episode (CCE), they estimated a median cost of 7,404.5 euros for the acute phase and a median cost of 3,449.6 euros for the rehabilitative one. Therefore, an effective fall prevention intervention is of primary importance also to reduce this tremendous socioeconomic burden.

A systematic literature review and meta-analysis analyzed fall risk factors in community-dwelling older people (6), highlighting that falling results from an interaction between environmental hazards and inadequate physiology to cope with them, such as gait problems, poor vision, impaired peripheral sensation, lower limb strength, dizziness, and the use of psychotropic medications or polypharmacy (6, 7). In older adults presenting for medical attention after a fall or who have gait or balance problems, guidelines recommend a multifactorial fall risk assessment (8). This strategy implies identifying modifiable risk factors and implementing targeted interventions for fall prevention (3). However, the delivery of effective treatments for fall prevention in usual clinical practice still represents a challenge (9, 10).

According to a recent Cochrane review (1) on fall prevention for older people living in the community, three effective interventions were identified: single, multiple, and multifactorial interventions. This systematic review identified that both multifactorial intervention and exercise alone, either delivered as a multiple-component group exercise or home-based exercise, reduce fall rates but only exercise reduces fall risk. Multifactorial programs also effectively reduced falls, even though trials on this intervention are heterogeneous (11). In a recent ongoing Randomized Control Trial (RCT) (12), a combination of multiple and multifactorial interventions was employed to prevent falls in community-dwelling older people. However, results on treatment effectiveness are not available yet. In 2020 Lamb et al. demonstrated that screening by mail followed by a targeted exercise intervention or multifactorial approach to preventing falls did not result in a lower rate of fractures than advice by mail alone (13). Moreover, RCTs in the Cochrane systematic review did not include subjects with neurological conditions, such as Parkinson’s Disease (PD) and stroke.

Evidence from the literature showed that among people affected by these neurological conditions, a high proportion of fallers is recorded together with a high rate of participation restriction (14, 15). Previous studies suggested that exercise can improve balance in PD, even though the fall rate and fall risk reduction were not achieved (16–18). A recent study (19) investigated a combination of educational and exercise interventions to reduce falls in people with neurological conditions: results from this RCT did not show a reduction in fall risk. However, to the best of the authors’ knowledge, no studies were conducted on a combined intervention to prevent falls in the elderly living in the community, including participants affected by neurological conditions, and with a synergy between group exercise and personalized home exercise to increase compliance and chances that home exercise becomes an integral part of a long-term more active and healthier lifestyle.

Thus, the current study aimed to evaluate the efficacy of a multiple-component intervention associated with a personalized multifactorial intervention, to reduce fall rates in community-dwelling older adults who can walk but are at risk of falling, including those with PD and stroke, compared to usual care. We hypothesized that the intervention group would present a lower number of falls, a lower fall probability, and a longer time to the first fall at a twelve-month follow-up than the control group.

## 2 Materials and methods

### 2.1 Study design

This study was a multicenter randomized controlled trial where individuals randomized to the treatment group (TG) received an 11-week multiple-component and personalized multifactorial intervention to reduce fall risk, whereas participants in the control group (CG) received only usual care. Pre-test and post-test assessments were conducted, respectively, before randomization and twelve weeks after the commencement of the intervention. Primary and secondary endpoints were assessed at a twelve-month follow-up. The study design is presented in Figure 1.

**Figure 1.**
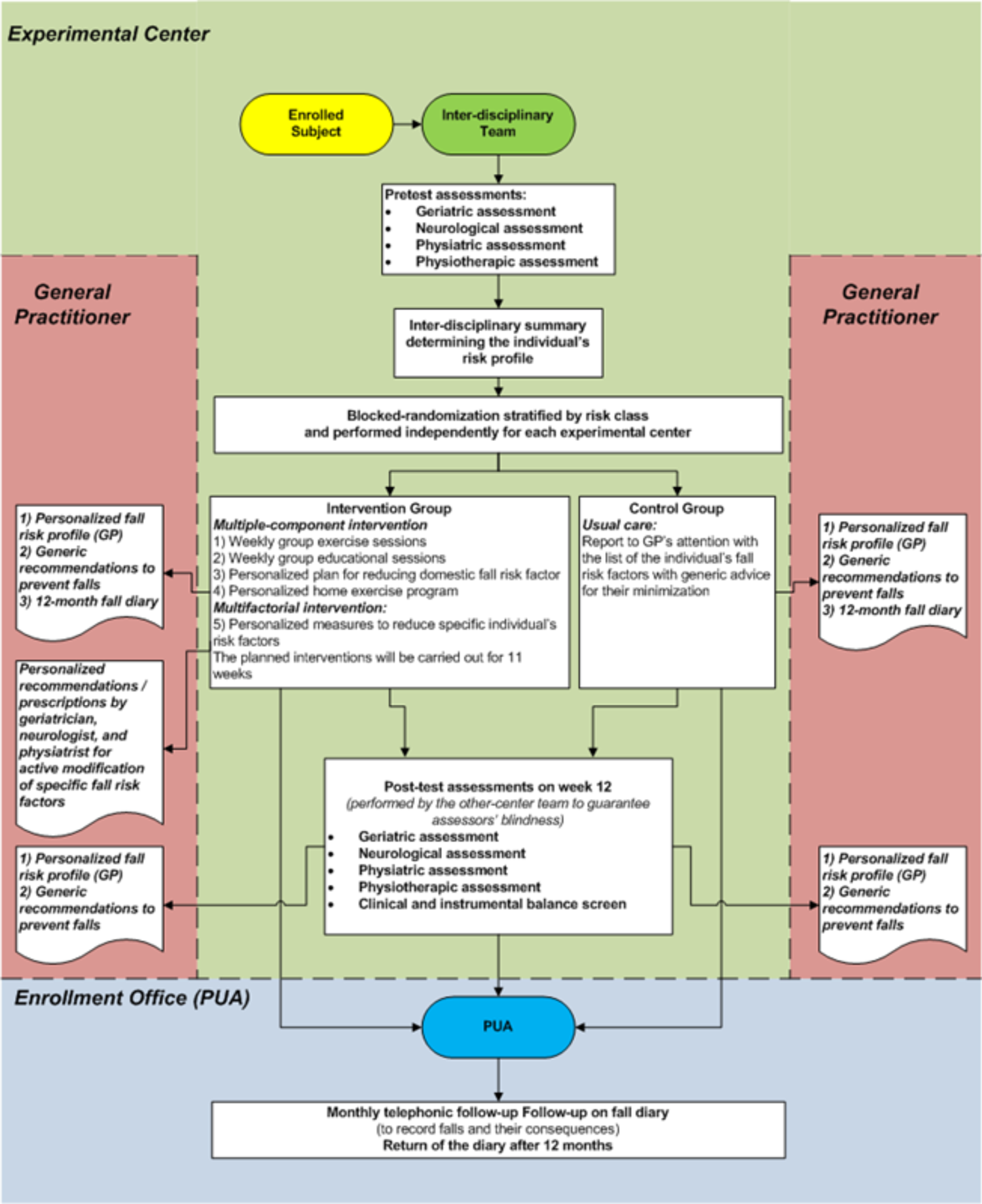
Study summary flow chart

The study was conducted in two Italian Public Hospitals (Ospedale Civile di Baggiovara in Modena and Arcispedale Santa Maria Nuova in Reggio Emilia) between 2015 and 2016. It was registered on clinicaltrials.gov (unique identifier NCT03592420) and approved by the local Ethical Committee (Provincial Ethics Committee of Modena 1141/CE/2014). Furthermore, all participants gave written informed consent to participate in the study, which was conducted in strict adherence to the ethical principles of the Helsinki Declaration (19).

### 2.2 Participants

Inclusion criteria were: age ≥65 years, moderate-to-high fall risk associated with age and/or neurological conditions (i.e., PD and stroke), ability to walk for at least 10 meters without assistance (possible use of a walking aid), and agreement to give written informed consent to the study.

Exclusion criteria were: severe general health conditions interfering with physical exercise, cognitive impairment (Mini-Mental Test score <24 or cognitive conditions interfering with test administration), severe deafness, severe vision impairment, severe aphasia or visual-spatial disorders, subjective and objective vertigo in the last three months, ongoing physiotherapy likely to influence the target variables (at the time of enrolment).

### 2.3 Enrolment algorithm

Health professionals (medical specialists or general practitioners) could signal to an enrolment office shared between the two centers (‘Punto Unico di Arruolamento’, PUA) through an ad hoc case report any subjects over 65 years old, with or without a diagnosis of PD or stroke, considered to be ‘at fall risk’. The subject’s compliance with the inclusion and exclusion criteria was declared in this form. Also, subjects considering themselves at risk of falling could self-report themselves through a dedicated email address.

After the initial report, potentially eligible participants were screened for inclusion and exclusion criteria in more detail. In particular, eligible subjects underwent further assessments by these subsequential steps:

1. Anamnestic Assessment of Eligibility (AAE):
  - This assessment was the PRE.C.I.S.A. first selection step. It was administered by a PUA’s trained nurse, through a telephone call, to older adults who had been signaled as ‘at fall risk’;
  - The aim was to confirm the inclusion/exclusion criteria for the study recruitment and evaluate the most influential fall risk factors. In particular, the PUA’s nurse verified the subject’s adherence to inclusion and exclusion criteria. Furthermore, the subject was submitted to the Fall Risk Assessment Tool (FRAT, Nandy, 2004) (20), was asked if he/she was afraid to fall, and it was verified his/her ability to walk 10 meters without assistance;
  - Later, each subject was classified as ‘not eligible and/or at low risk’, ‘moderate’, or ‘high’ fall risk according to the defined algorithm in Table 1;
  - Those who resulted at ‘low fall risk’ or ‘not satisfying study criteria’ were excluded. However, the study protocol allowed the PUA’s nurse to contact these subjects after one year to record any eventual fall, thus providing helpful quantitative information for the post-hoc validation of the screening algorithm.
  - After combining the assessment results, people who resulted at ‘moderate-to-high fall risk’ accessed the successive selection step (Objective Assessment of Eligibility - OAE).
  2. Objective Assessment of Eligibility (OAE):
    - This assessment constituted the PRE.C.I.S.A. second selection step, and it was administered during an outpatient visit by a trained physiotherapist to individuals selected at ‘moderate-to-high fall risk’ during the previous selection step (AAE);
    - The aim was to evaluate in detail all fall risk factors described in the literature and, hence, to confirm the eligibility for the study (be at ‘moderate-to-high fall risk’ after the combination of the assessment results). In particular, the prospective participant was submitted to the Falls Risk for Older People in the Community Screen (FROP-Com Screen) (21), to the Fall Risk Assessment Tool (FRAT, Stapleton, 2009) (22), to several mobility and balance tests (10 Meters Walking Test, Timed Up&Go test, Tandem stance from the 4 Stage Balance Test, 30-second Chair Standing test, Short Physical Performance Battery, Functional Reach Test), to the Abbreviated Mental Test Score, and the visual acuity assessment (Snellen Chart);
    - Those who obtained an ‘eligible coefficient’ ≥1, calculated from the FROP-Com Screen and the FRAT (Stapleton, 2009) total scores, were judged as ‘eligible’ to be enrolled in the study (Table 1);
    - Any individual re-classified at ‘low risk’ following this second assessment step was excluded from the enrolment and contacted one year later by the PUA’s trained nurse to collect the number of any eventual falls that occurred. This last contact allowed us to collect quantitative information for the post-hoc validation of the screening algorithm.

**Table 1.** AAE and OAE enrolment algorithms

| Anamnestic assessment of eligibility (AAE) algorithm | Objective assessment of eligibility (OAE) algorithm |
| --- | --- |
| *Not eligible and/or at low risk*<br>the absence of at least one inclusion criteria OR<br>the presence of at least one exclusion criteria OR the inability to walk 10 meters without assistance OR<br>the ability to walk 10 meters without assistance AND a FRAT (Nandy, 2004) total score equal to 0 AND the absence of fear of falling. | *FROP-Com Screen*<br>Total score 0-1: low risk (eligible coefficient = 0)<br>Total score 2-4: medium risk (eligible coefficient = 1)<br>Total score 5-9: high risk (eligible coefficient = 2)<br>*FRAT (Stapleton, 2009)*<br>Total score 5-11: low risk (eligible coefficient = 0)<br>Total score 12-15: medium risk (eligible coefficient = 1)<br>Total score 16-20: high risk (eligible coefficient = 2). |
| *Moderate risk*<br>the ability to walk 10 meters without assistance AND the absence of previous fall(s) AND<br>a FRAT (Nandy, 2004) total score $\leq 2$ OR<br>the presence of fear of falling. |  |
| *High risk*<br>the ability to walk 10 meters without assistance AND<br>the presence of previous fall(s) OR<br>a FRAT (Nandy, 2004) total score $\geq 3$. |  |

### 2.4 Randomization

After the enrolment, subjects judged as ‘eligible’ participants underwent the pre-test assessments, which were conducted by a Physiatrist (P), a Physiotherapist (PT), a Geriatrician (G), and a Neurologist (N), as described in the following outcome measures section. These assessments helped determine in detail the individual fall risk profile.

Subsequently, the last assessor, who performed the pre-test evaluation, randomized the enrolled subject to the intervention group (IG) or the control group (CG). A web-based database was developed ad hoc for this study, and computer-based randomization was implemented to guarantee the allocation concealment. The randomization sequence was created using random block sizes of 4. Participants were stratified by risk classes (older adults 65-79 years without associated neurological disease; older adults ≥80 years without associated neurological disease; older adults with stroke; older adults with PD) and independently for each center.

After the randomization, all enrolled subjects were informed about the allocation arm and received a ‘usual care’ intervention (figure 1) based on:

- a report on their individual risk factor profile;
- an illustrated brochure on fall prevention;
- personalized suggestions to minimize the fall risk addressed to their General Practitioner (GP).

Furthermore, all participants were provided with a one-year fall report diary, integrated with a physical activity monitoring diary, and several copies of a ‘fall report’ that had to be filled by the participant, in case of a fall, with more detailed information about the event.

### 2.5 Interventions

Participants in the IG were taken in charge by an interdisciplinary team, including the four professionals mentioned above, who administered synergically the following five interventions (described in detail in Appendix 1).

#### 2.5.1 Group exercise sessions

This intervention was based on eleven weekly group sessions (including six participants) for sixty minutes. Each session was composed of the following parts:

- Warming-up (five minutes): head, neck, trunk, and ankle movements, back and knee extensions, walking on the spot;
- Circuit training (thirty-five minutes): muscular strength exercises, balance exercises, and recovery techniques from falling (23);
- Dynamic balance and walking exercises specific for the risk class (ten minutes). The remaining ten minutes were used to check the physical activity report diary.

In the first session, the PT delivered a weight vest to each IG participant and verified the initial level for each cited three circuit training station.

At each group session, the participant needed to have his/her weight vest, his/her fall-physical activity report diary of the current month, and his/her manual of the home exercise program (see next point 4). During rest periods from exercises, the participant delivered his/her fall-physical activity report diary and, in case of at least one fall during the week, the completed ‘fall report’. At the end of the session, the PT updated the manual of the home exercise program with the week level progression of the exercises (the passage changed every two weeks, but depending on individual need, it was possible to add other series of the same exercise in the intermediate weeks).

#### 2.5.2 Group education sessions on fall risk factors

The IG participants received a thirty-minute educational session after each weekly group exercise session, focussing on different modifiable fall risk factors or avoidable risky behavior. The educational session was divided into two parts:

i. a ten-minute frontal lecture on a specific theme held by a component of the interdisciplinary team;
ii. a twenty-minute group discussion on the lecture content (involving participants, caregivers, and professionals). A handbook summarizing these topics was provided to each participant at the beginning of the first education session.

#### 2.5.3 Personalized plan for reducing domestic fall risk factors

During the first week of treatment, a PT performed a home visit for each IG participant. During this visit, usually lasting sixty to ninety minutes, the PT:

- Filled the ‘Home environmental risks questionnaire’ and compared it with the same questionnaire compiled by the participant at the pre-test assessment;
- Gave specific recommendations with proposals for correcting the detected modifiable risk factors by delivering the ‘Suggestions for the reduction of environmental risks at home’ information sheet where the actual hazards were highlighted;
- Verified the presence of the fall-physical activity report diary, delivered at the time of recruitment, in a position that facilitated its compilation in case of fall (e.g., hanging on the wall in the living room/ kitchen, etc.);
- During the subsequent three home visits related to the personalized home exercise program, the PT checked the implementation of the recommended interventions proposed during the first home access and filled the ‘Check-list of correction of environmental risk factors at home’.

#### 2.5.4 Personalized home exercise program

This intervention was coordinated with the group exercise program aimed at improving strength, and static and dynamic balance, with the specific aim of enabling the participants to develop a long-term daily habit of exercising and performing physical activity in the context of a progressive and permanent adoption of a healthy and active lifestyle.

- The PT devised this program in the context of an initial home visit (on the second week) and subsequently monitored within two further home visits (on the fourth and sixth weeks).
- During the initial home visit, an illustrated manual containing strength and balance exercises was provided and explained to each participant based on the first group exercise session. These exercises were chosen between those the subject performed with greater safety in the group session.
- Besides, the PT gave indications about recommended training frequency and time and registration of the performed physical activity in his/her fall-physical activity report diary of the current month.
- During the subsequent two visits, the PT verified: a) the setting adequacy, b) the modality in which the participant performed the suggested exercises, and c) the update of the fall-physical activity report diary.
- Finally, in all three home accesses linked to the home exercise program, the PT checked the implementation/maintenance of the recommendations on risk factors correction given in the first-week home visit.

#### 2.5.5 Multifactorial personalized intervention

This intervention aimed at modifying additional risk factors, which were performed by the interdisciplinary team and included the following interventions:

- Review of medications, including psychotropic medications (N and G), antiparkinsonian drugs (N), and cardiovascular medications (G);
- Management of unaddressed visual impairments (G): ophthalmologist referral, lens prescription, suggestions regarding the limitation of bifocal lenses;
- Management of unaddressed cardiovascular issues (G), such as postural hypotension, covert cardiac failure, and abnormalities of cardiac rhythm, eventual cardiology referral;
- Vitamin D prescription (G);
- Improvement of nutritional state (G), with prescription of caloric-proteic integration and/or nutritional referral;
- Management of muscle-skeletal issues, including spasticity (P and PT);
- Education about foot self-care, including podologist referral if appropriate (P);
- Assessment, prescription, and final testing of orthosis and mobility aids, including proper shoes (P and PT).

#### 2.5.6 Interventions delivery

Interventions one to four were administered to all IG participants (multiple-component intervention), whereas the multifactorial intervention (intervention number five) was personalized based on the individual fall risk profile devised on the pre-test assessment. Furthermore, interventions one, two, and five were conducted within an outpatient setting, while interventions three and four were home-based.

### 2.6 Comparator

Participants allocated to the CG received only the usual care, as described in the randomization section. The management of the fall risk of each individual enrolled in the CG was delegated to the participant’s GP.

### 2.7 Outcome measures

Participants’ demographic and clinical characteristics were collected during the baseline pre-test visit, including age, sex, fall risk according to epidemiological criteria, and Falls Risk for Older People in the Community (FROP-Com) criteria.

Further, several indicators were used to assess functioning at pre-test (T1) and three-month follow-up (T3), linkable to the International Classification of Functioning (ICF(24)) domains (body functions, activity and participation, environmental factors). In addition, even instruments administered at the OAE assessment were recollected at the three-month follow-up (T3) (see Appendix 2 for details).

The primary endpoint was represented by the total number of falls in each group over twelve months. A fall was defined as an ‘unexpected event where a person inadvertently comes to rest on the ground, floor, or lower level’ (25, 26). The secondary endpoints were other fall-related indicators (fall rate of subjects with one or more falls, fall severity, fall probability, and time to the first fall) recorded at the twelve-month follow-up.

Each participant was provided with their own fall diary and was followed up for twelve months with monthly telephone contacts to record the primary and secondary endpoints. During these monthly calls, each participant was inquired about any incurred falls at each contact, with date, circumstances, underlying cause, and related injuries. The primary endpoint was further verified at the end of the study by returning the fall diary.

The blindness of the assessments was guaranteed with various strategies:

- For both pre-test and post-test evaluations, as the former was performed before randomization, whereas the latter by the other center’s assessors, unaware of the allocation arms of the participants’ within the enrolling center;
- Furthermore, subjects in both groups were instructed not to discuss their allocation with other participants and assessors during the post-test assessments;
- Finally, the assessor was unaware of the allocation arms at the monthly follow-up calls.

### 2.8 Statistical Analyses

#### 2.8.1 Sample size calculation

The sample size was determined based on the assumptions that the fall risk in the control group was equal to 50% and that the experimental intervention was able to reduce this risk by 30%, that is, to obtain a fall risk in the treatment group equal to 35%. Fixing the type I error at 0.05 (95% confidence level) and the type II error at 0.20 (80% power), it was decided to enlist at least 366 subjects (183 per group).

#### 2.8.2 Descriptive statistics for all participants

Descriptive statistics were calculated at the time of enrolment in the study. Summary statistics were means and standard deviations for quantitative variables, median and interquartile ranges for categorical variables, and absolute frequencies and percentages for nominal variables.

#### 2.8.3 Primary and secondary endpoint calculations

The number of falls recorded monthly by telephone interview was the basic element for the primary endpoint calculation. In particular, the monthly fall number was added for all twelve months of follow-up to obtain the number of falls observed during the entire period of inclusion in the study of each participant.

The start and end date of the follow-up were needed to calculate the time to the first fall (secondary endpoint). Therefore, the date of randomization for each subject was considered the start date for the follow-up. The end date of the follow-up was calculated differently for participants with at least one fall and those without falls. For the former, we considered the least recent date among the dates of telephone interviews in which at least one fall was reported. For the latter, the most recent date among telephone interviews was considered. Thus, the follow-up time in months was equal to the difference in days between the start and end follow-up dates, divided by 30.4 (mean duration of a month).

#### 2.8.4 Analysis of the differences between groups (IG and CG)

Concerning the study’s primary endpoint, the comparison of observed fall incidences in the two groups was evaluated using statistical regression methodologies for counting data. A model assumes a negative binomial distribution for the response variable (number of falls). The results were expressed as the Incidence Rate Ratio (IRR) with a 95% confidence interval (95CI%) and a p-value, comparing the experimental and control groups.

Concerning the secondary endpoint ‘fall probability’, the results were expressed as Relative Risk (RR) with a 95%CI and a p-value, referring to the comparison between IG and CG. This fall probability was calculated for one fall, two or more falls, and three or more falls (multiple fallers) within twelve months.

A Cox regression model performed analyses of the secondary endpoint ‘time to the first fall’. The results were expressed as Hazard Ratio (HR) with a 95%CI and a p-value, comparing the two groups. In addition, the cumulative probabilities of occurrence of at least one fall were graphically represented as Kaplan-Meier survival curves, reporting the survival point estimate from falls at three, six, and twelve months, with 95%CI.

#### 2.8.5 Analysis of the differences between sub-groups (four etiological risk class categories)

All assessments of the observed differences between randomization arms (IG and CG) were repeated separately in the four subgroups identified by the four etiological risk class categories considered in the study: age between 65 and 80, age over 80, elderly with Parkinson’s Disease, elderly with a previous stroke.

#### 2.8.6 Analysis of the differences between groups (IG and CG) for T3 endpoints (post-test)

##### 2.8.6.1 Rasch analysis

Preliminary to comparing the two groups on post-test with ANCOVA, we performed a Rasch analysis of the scale and questionnaires involved in the comparison. Rasch analysis was conducted because ANCOVA is a parametric statistical analysis requiring continuous variables, whereas the total scores of scales and questionnaires deliver ordinal data. Indeed, within Rasch analysis, it may be possible to transform the ordinal total score of a scale or a questionnaire into interval-level person estimates of ability, should the data fit the requirement of the Rasch model (i.e., the mathematical model upon which Rasch analysis relies) (27). In particular, the Rasch analysis focused on the following indicators:

- FROP-Com (28);
- Berg Balance Scale (BBS (29, 30));
- Performance-Oriented Mobility Assessment (POMA (30, 31));
- Fullerton Advanced Balance Scale (FABS (30, 32));
- Mini-BESTest (33).

The FROP-Com is a global fall risk indicator, while the other four indicators all quantify balance, although with differences related to the measurement range. Therefore, two Rasch analyses were carried out separately: the first for the FROP-Com and the second for the four balance indicators. Given their conceptual equivalence (30), items of single balance scales were treated as testlets in the latter analysis. Testlets (or super-items) are sum scores from a set of associated items. Thus, the Rasch analysis was conducted on four testlets, one for each balance scale (34). This approach was adopted to absorb the local dependence between the items of the various balance scales (30,34–36).

##### 2.8.6.2 Pre-test vs. post-test differences between groups (IG and CG)

The values of the above five indicators, calculated before and after the intervention, were compared between the two groups using parametric statistical techniques. In particular, we reported the mean values of these parameters at the pre-test and post-test levels. The post-intervention values were compared between the groups through a linear regression model that uses the treatment and the pre-intervention value as independent variables (ANCOVA model). This analysis was reported as mean differences (MD) with 95% and p-value confidence interval. The effect size was calculated as the Cohen’s d by comparing pre-post differences between groups.

#### 2.8.7 Cases lost to follow-up

Whenever possible, the reasons for any cases lost to follow-up were recorded. Concerning logistic regression, an analysis that considered all randomized subjects without considering any follow-up loss was initially conducted according to the principle of the intention to treat. In case of loss to follow-up due to death or other causes, information collected up to that time was considered. Should a subject be lost to follow-up, independently from experiencing a fall or not (primary outcome), his/her data were considered for the analysis. Thus, it was possible to conduct sensitivity analyses that hypothesized various scenarios of the outcomes considered for loss to follow-up.

Regarding the analysis of survival curves, any loss to follow-up data was treated as censored since the last available information for these subjects. However, it was possible to conduct further sensitivity analyses even in this context.

#### 2.8.8 Statistical software

Statistical analyses were performed using the Stata 14 software (StataCorp LP, College Station) and R 3.4.3 (the R Foundation for Statistical Computing, Wien) by the Medical Statistics Unit of the University of Modena e Reggio Emilia, using a 95% confidence level (p 0.05). In addition, Rasch analyses were carried out using the software RUMM 2030 (version 5.4 for Windows. RUMM Laboratory Pty Ltd, Perth, Australia: 1997-2017; www.rummlab.com).

## 3 Results

### 3.1 Descriptive statistics for all participants (n=403)

Seven hundred ninety-one participants were assessed for eligibility, and four hundred and three were included in the study and randomized to either the CG (n=200) or the IG (n=203) (table 2 and figure 2). Seventy-one subjects (forty-eight in the CG and twenty-three in the IG group) were lost to follow-up (figure 2); between them, respectively, fifteen and eleven elderly people interrupted their participation during the treatment period (‘discontinued intervention’, figure 2). The two centers enrolled almost an equal number of patients (49.1% and 50.9% at Modena and Reggio Emilia, respectively).

**Figure 2.**
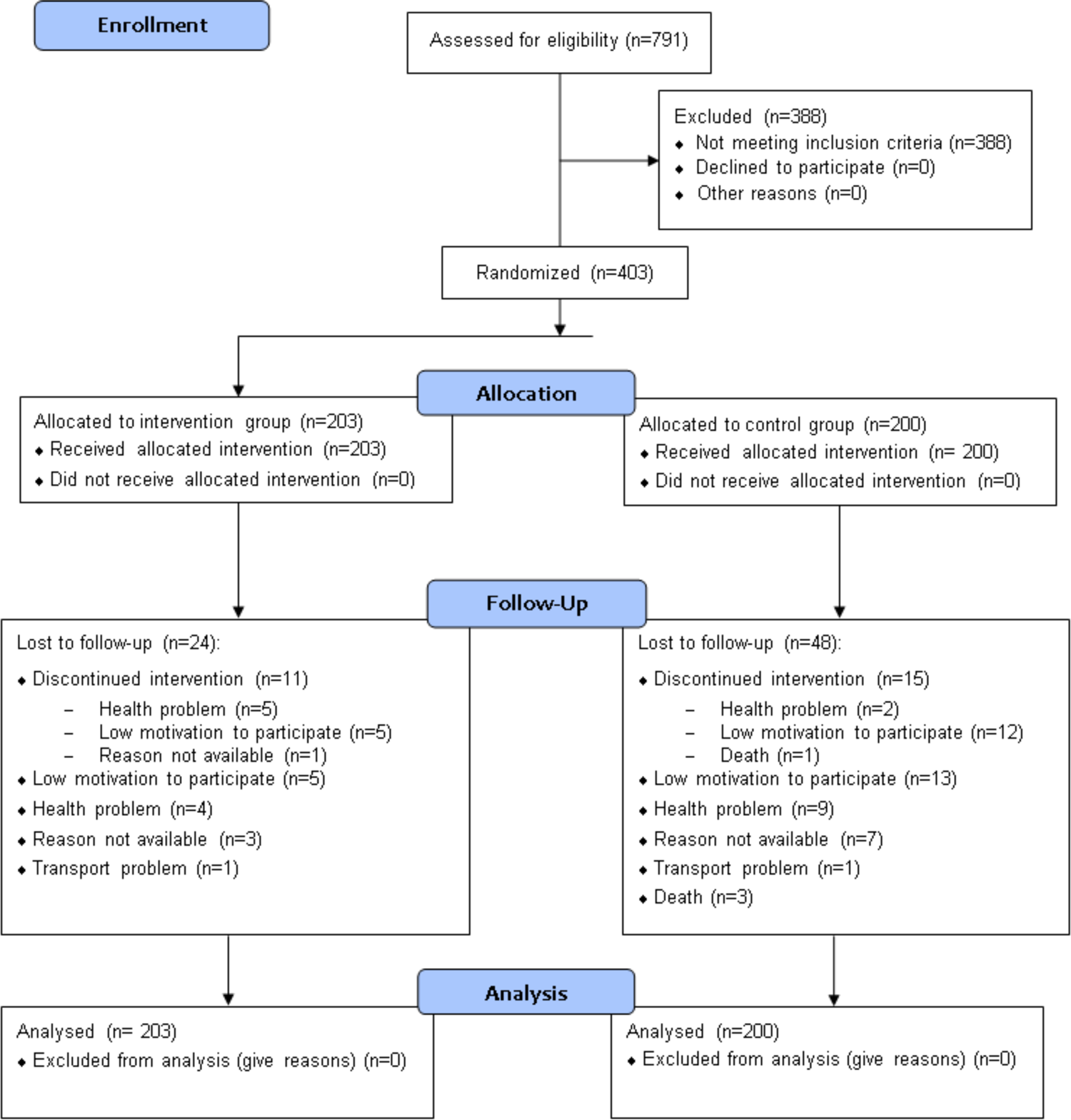
CONSORT 2010 flow diagram

**Table 2.**
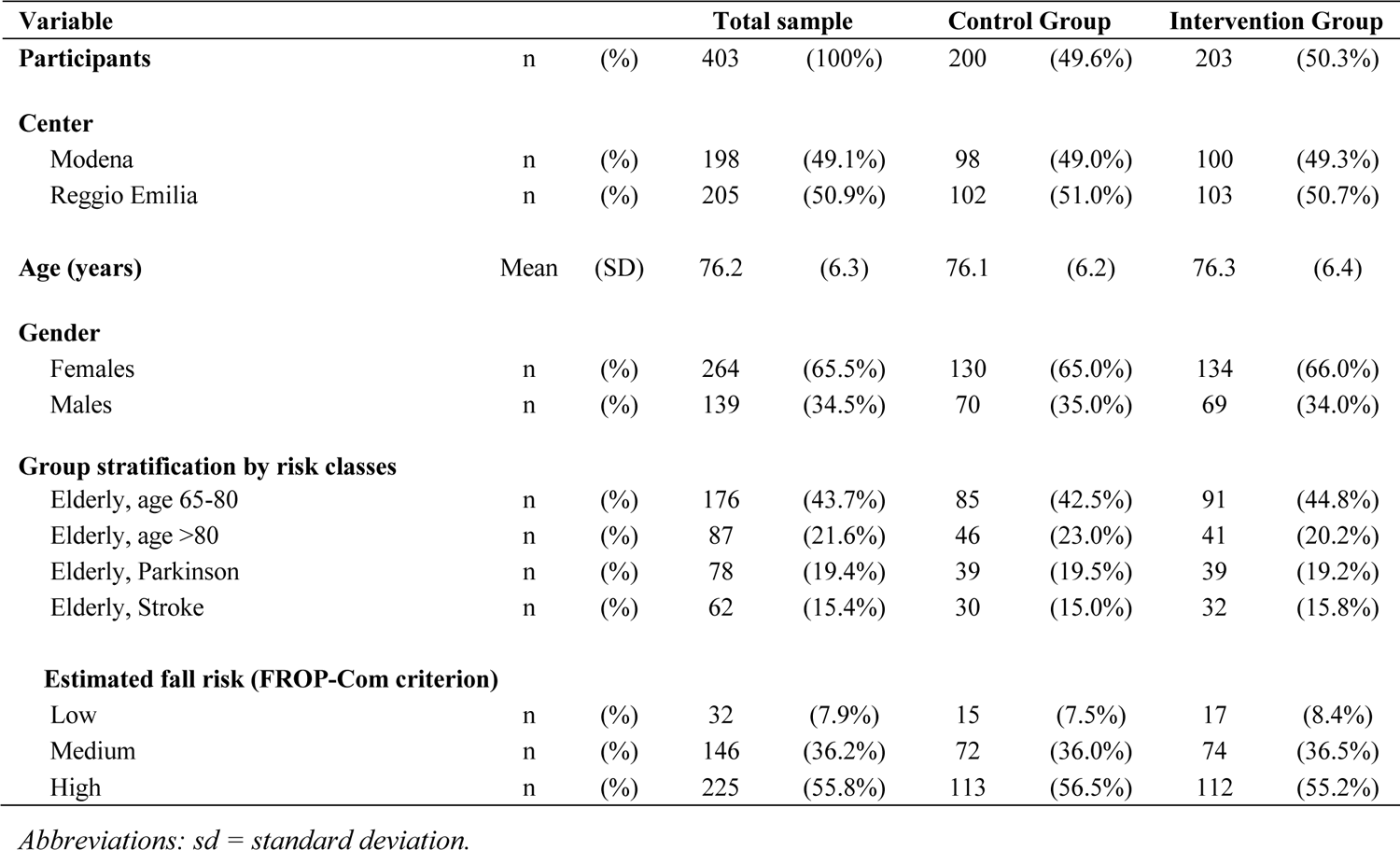
Clinical and demographic sample characteristics

| Variable |  |  | Total sample |  | Control Group |  | Intervention Group |  |
| --- | --- | --- | --- | --- | --- | --- | --- | --- |
| Participants | n | (%) | 403 | (100%) | 200 | (49.6%) | 203 | (50.3%) |
| **Center** |  |  |  |  |  |  |  |  |
| Modena | n | (%) | 198 | (49.1%) | 98 | (49.0%) | 100 | (49.3%) |
| Reggio Emilia | n | (%) | 205 | (50.9%) | 102 | (51.0%) | 103 | (50.7%) |
| Age (years) | Mean | (SD) | 76.2 | (6.3) | 76.1 | (6.2) | 76.3 | (6.4) |
| **Gender** |  |  |  |  |  |  |  |  |
| Females | n | (%) | 264 | (65.5%) | 130 | (65.0%) | 134 | (66.0%) |
| Males | n | (%) | 139 | (34.5%) | 70 | (35.0%) | 69 | (34.0%) |
| **Group stratification by risk classes** |  |  |  |  |  |  |  |  |
| Elderly, age 65-80 | n | (%) | 176 | (43.7%) | 85 | (42.5%) | 91 | (44.8%) |
| Elderly, age >80 | n | (%) | 87 | (21.6%) | 46 | (23.0%) | 41 | (20.2%) |
| Elderly, Parkinson | n | (%) | 78 | (19.4%) | 39 | (19.5%) | 39 | (19.2%) |
| Elderly, Stroke | n | (%) | 62 | (15.4%) | 30 | (15.0%) | 32 | (15.8%) |
| **Estimated fall risk (FROP-Com criterion)** |  |  |  |  |  |  |  |  |
| Low | n | (%) | 32 | (7.9%) | 15 | (7.5%) | 17 | (8.4%) |
| Medium | n | (%) | 146 | (36.2%) | 72 | (36.0%) | 74 | (36.5%) |
| High | n | (%) | 225 | (55.8%) | 113 | (56.5%) | 112 | (55.2%) |

*Abbreviations: sd = standard deviation.*

The mean age of enrolled participants was 76.2 years (SD: 6.3), and about two-thirds of them (65.5%) were females. About two-thirds of the enrolled patients (65.2%) were elderly with an estimated fall risk at one year comprised between 33% and 50%, as 43.7% and 21.6% were classified within the 65 to 80 and >80 risk classes, respectively. The remaining 34.7% were elderly patients with an associated neurological condition. Their estimated fall risk at one year was between 60% and 70%, as 19.4% and 15.4% of them had a diagnosis of Parkinson’s Disease or stroke, respectively. Considering the estimated fall risk of the enrollment patients, only 7.9% could be considered at ‘low risk’ according to the FROP-Com, whereas the enrollment algorithm classified the remaining 92.1% of patients correctly as being at moderate (36.2%) or high fall risk (55.8%) according to the FROP-Com. The subjects’ characteristics at baseline were balanced between the two groups.

Amongst the participants, the majority of them (58.6%) experienced at least one or more falls. In particular, 21.8% and 14.6% experienced one or two falls, respectively, whereas the percentage of multiple fallers (>2 falls) was 22.1% (table 3). The rate of fallers, defined as those with at least two falls, was about one-third (36.7%). Regarding the primary endpoint, six hundred ninety falls were reported at the twelve-month follow-up (table 3), with a mean number of falls per participant equal to 1.71 (SD 3.36).

**Table 3.** Endpoint evaluation between groups (IG and CG)

|  |  |  | Total |  | Control Group |  | Intervention Group |  |
| --- | --- | --- | --- | --- | --- | --- | --- | --- |
| Participants | n | (%) | 403 | - | 200 |  | 203 |  |
| **Participants by falls (no falls vs. ≥1 falls)** |  |  |  |  |  |  |  |  |
| 0 falls | n | (%) | 167 | (41.4%) | 83 | (41.5%) | 84 | (41.4%) |
| ≥1 falls | n | (%) | 236 | (58.6%) | 117 | (58.5%) | 119 | (58.6%) |
| One fall | n | (%) | 88 | (21.8%) | 40 | (20.0%) | 48 | (23.6%) |
| Two falls | n | (%) | 59 | (14.6%) | 26 | (13.0%) | 33 | (16.3%) |
| More than two falls (multiple fallers) | n | (%) | 89 | (22.1%) | 51 | (25.5%) | 38 | (18.7%) |
| **Participants by falls (0-1 falls vs. ≥2 falls)** |  |  |  |  |  |  |  |  |
| No fallers (0-1 falls) | n | (%) | 255 | (63.3%) | 123 | (61.5%) | 132 | (65.0%) |
| Fallers (≥2 falls) | n | (%) | 148 | (36.7%) | 77 | (38.5%) | 71 | (35.0%) |
| **Fall rate (primary endpoint)** |  |  |  |  |  |  |  |  |
| Total number of falls | n | (%) | 690 | - | 353 | (51.2%) | 337 | (48.8%) |
| Mean number of falls per participant | Mean | (SD) | 1.71 | (3.33) | 1.77 | (3.17) | 1.66 | (3.49) |
| **Falls by injury** |  |  |  |  |  |  |  |  |
| No injury | n | (%) | 465 | (67.4%) | 238 | (67.4%) | 227 | (67.4%) |
| Minor injury, no medical consultation | n | (%) | 106 | (15.4%) | 57 | (16.2%) | 49 | (14.5%) |
| Minor injury, with medical consultation | n | (%) | 75 | (10.9%) | 34 | (9.6%) | 41 | (12.2%) |
| Serious injury | n | (%) | 44 | (6.4%) | 24 | (6.8%) | 20 | (5.9%) |
| **Probability of absence of falls (Kaplan-Meier)** |  |  |  |  |  |  |  |  |
| Time to the first fall in months | Median | (95%CI) | 11.1 | (9.4-12.3) | 11.1 | (7.6-12.3) | 11.2 | (9.7-NA) |
| at three months | % | (95%CI) | 77.6% | (73.7-81.8%) | 76.5% | (70.8-82.6%) | 78.8% | (73.4-84.6%) |
| at six months | % | (95%CI) | 65.0% | (60.5-69.8%) | 63.4% | (57.0-70.4%) | 66.5% | (60.3-73.3%) |
| at twelve months | % | (95%CI) | 47.3% | (42.6-52.4%) | 46.8% | (40.3-54.2%) | 47.8% | (41.4-55.2%) |

*Abbreviations: sd = standard deviation; CI95% = confidence interval at 95% level; NA = not applicable.*

Most falls (67.4%) led to no injury, whereas 32.6% were associated with various injuries. In particular, 6.4% of them led to serious injury requiring hospitalization, whereas 10.9% and 15.4% of falls were associated with minor injury requiring or not a medical consultation, respectively.

The median time of occurrence of the first fall was 11.1 months. A probability of absence of falls of 77.6% (95%CI [73.7, 81.8%]), 65.0% (95%CI [60.5, 69.8%]) and 47.3% (95%CI [42.6, 52.4%]) were recorded, respectively, at three, six and twelve months.

### 3.2 Differences between groups (CG and IG)

In the CG and the IG, most participants (58.5% and 58.6%, respectively) fell at least one or more times. In particular, 20.0% and 23.6% experienced one fall, 13.0% and 16.3% two falls, whereas the percentage of multiple fallers (>2 falls) was 25.5% and 18.7%, respectively (table 3). The percentage of fallers with at least two falls was 38.5% and 35% in the CG and the IG, respectively. Regarding the primary endpoint, 353 falls were reported at the twelve-month follow-up in CG, compared to 337 in the IG (table 3), with a mean number of falls per participant equal to 1.77 (SD 3.17) and 1.66 (SD 3.49) falls, respectively. Fall distribution by groups is reported in figure 3.

**Figure 3.**
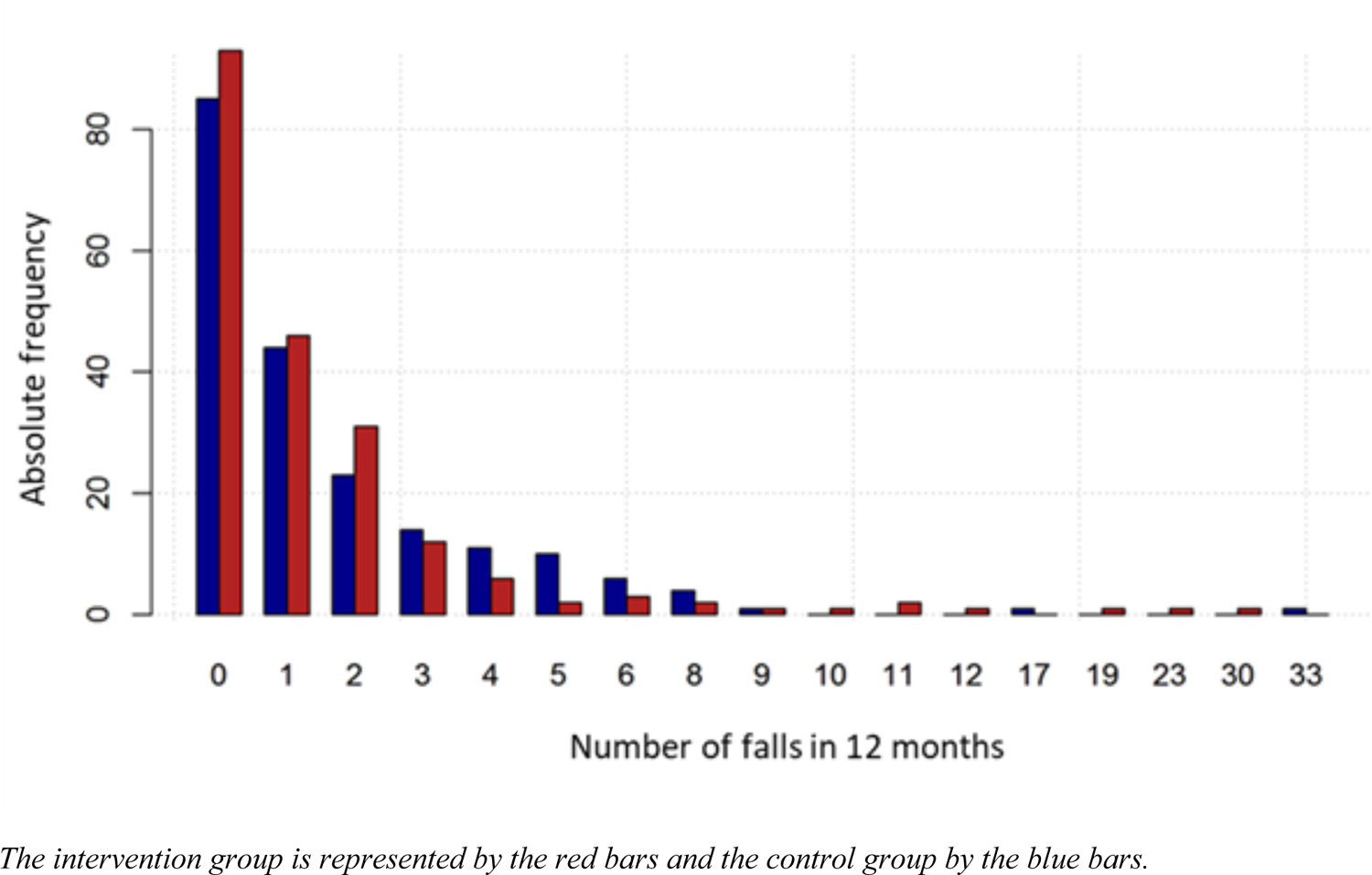
Fall number distribution by group

Most falls (67.4%) led to no injury for both groups, whereas the remaining one-third was associated with various degrees of injury. In particular, 16.2% in the CG and 14.5% in the IG of falls led to minor injuries not requiring medical consultation, whereas 9.6% and 12.2% of falls were associated with minor injuries requiring medical consultation. Finally, 6.8% of falls in the CG and 5.9% in the IG led to serious injury requiring hospitalization (table 3).

A probability of absence of falls of 76.5% (95%CI [70.8, 82.6%]) and 78.8% (95%CI [73.4, 84.6%]) in the first three months were recorded, respectively, for CG and IG. No statistically significant differences were observed between groups regarding the number of falls (Incidence Rate Ratio - IRR=0.94, 95%CI [0.69-1.29], p=0.693), and the fall probability for one fall, and two or more falls (Risk Ratio one fall - RR=0.94, 95%CI [0.79 - 1.12], p=0.503; Risk Ratio two or more falls - RR=0.89, 95%CI [0.67 - 1.17], p=0.398). Regarding the falling probability for three or more falls (multiple fallers), this was in favor of the IG, although slightly above the chosen level of statistical significance (Risk Ratio three or more falls - RR=0.68, 95%CI [0.45 - 1.01], p=0.052) (table 4).

**Table 4.**
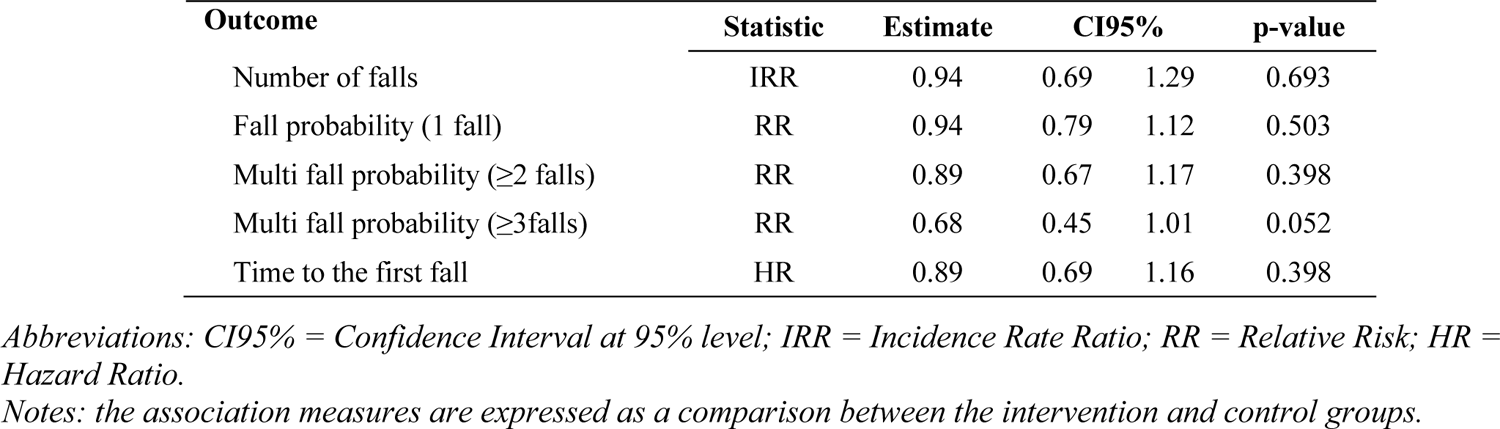
Analysis of observed differences between IG and CG

The median time to the first fall was 11.1 months (95%CI 7.6-12.3) in the CG and 11.2 months (95%CI 9.7-NA) in the CG. No statistically significant differences were observed between groups regarding the time to the first fall (Hazard Ratio - HR=0.89, 95%CI [0.69-1.16], p=0.398) (table 4).

### 3.3 Differences between sub-groups (four etiological risk class categories in the CG and IG)

Regarding the number of falls (table 5), the comparison between CG and IG showed a trend toward a lower (although not significant) risk of falling for elderly aged 65-80 (IRR=0.79, 95%CI [0.50, 1.25]), elderly aged >80 (IRR=0.85, 95%CI [0.51, 1.40]), and elderly with associated Parkinson’s Disease (IRR=0.94, 95%CI [0.52, 1.72]) in the IG. In addition, the risk appeared lower (although not significant) for the elderly with stroke sequelae (IRR=2.39, 95%CI [0.88, 6.49]) in the CG.

**Table 5.** Analysis of observed differences between subgroups in the IG and CG

| Outcome | Statistics | Estimate | 95CI% |  | p-value |
| --- | --- | --- | --- | --- | --- |
| Number of falls |  |  |  |  |  |
| Age 65-80 | IRR | 0.79 | 0.50 | 1.25 | 0.313 |
| Age >80 | IRR | 0.85 | 0.51 | 1.40 | 0.519 |
| Parkinson | IRR | 0.94 | 0.52 | 1.72 | 0.849 |
| Stroke | IRR | 2.39 | 0.88 | 6.49 | 0.086 |
| Fall probability (1 fall) |  |  |  |  |  |
| Age 65-80 | RR | 0.86 | 0.65 | 1.13 | 0.276 |
| Age >80 | RR | 1.03 | 0.74 | 1.47 | 0.829 |
| Parkinson | RR | 0.93 | 0.70 | 1.23 | 0.615 |
| Stroke | RR | 1.22 | 0.63 | 2.35 | 0.553 |
| Multi fall probability (≥2 falls) |  |  |  |  |  |
| Age 65-80 | RR | 0.72 | 0.33 | 1.55 | 0.397 |
| Age >80 | RR | 0.52 | 0.22 | 1.24 | 0.125 |
| Parkinson | RR | 0.62 | 0.36 | 1.05 | 0.068 |
| Stroke | RR | 3.75 | 0.44 | 31.7 | 0.185 |
| Multi fall probability (≥3 falls) |  |  |  |  |  |
| Age 65-80 | RR | 0.85 | 0.51 | 1.42 | 0.542 |
| Age >80 | RR | 1.21 | 0.65 | 1.95 | 0.682 |
| Parkinson | RR | 0.72 | 0.48 | 1.19 | 0.111 |
| Stroke | RR | 1.21 | 0.51 | 2.83 | 0.667 |
| Time to the first fall |  |  |  |  |  |
| Age 65-80 | HR | 0.83 | 0.56 | 1.25 | 0.381 |
| Age >80 | HR | 0.92 | 0.54 | 1.59 | 0.776 |
| Parkinson | HR | 0.83 | 0.49 | 1.41 | 0.497 |
| Stroke | HR | 1.32 | 0.58 | 3.01 | 0.509 |

*Abbreviations: CI95% = Confidence Interval at 95% level; IRR = Incidence Rate Ratio; RR = Relative Risk; HR =* *Hazard Ratio.*
*Notes: the association measures are expressed as a comparison between the four subgroups in the IG and CG.*

Concerning the falling probability for one fall, elderly aged 65-80 and with PD in the IG had a lower (although not significant) probability of falling than those randomized in the CG (RR=0.86, 95%CI [0.65, 1.13] and, respectively, RR=0.93, 95%CI [0.70, 1.23]). On the other hand, elderly aged >80 and elderly with stroke had a higher (although not significant) probability of falling if randomized in the IG (RR=1.03, 95%CI [0.74, 1.47] and, respectively, RR=1.22, 95%CI [0.63, 2.35]).

Concerning the falling probability for two or more falls, elderly aged 65-80 and aged >80 in the IG had a lower (although not significant) probability of falling than those randomized in the CG (RR=0.72, 95%CI [0.33, 1.55] and, respectively, RR=0.52, 95%CI [0.22, 1.24]). Even elderly with PD in the IG had a lower (although not significant) probability of falling than those randomized in the CG, with a RR closer to the chosen level of statistical significance (RR=0.62, 95%CI [0.36, 1.05]). Differently, the elderly with stroke had a higher (although not significant) probability of falling if randomized in the IG (RR=3.75, 95%CI [0.44, 31.7]).

Regarding the falling probability for three or more falls (multiple fallers), similarly to the probability for one fall, elderly aged 65-80 and with PD in the IG had a lower (although not significant) probability of falling than those randomized in the CG (RR=0.85, 95%CI [0.51, 1.42] and, respectively, RR=0.72, 95%CI [0.48, 1.19]). On the other hand, elderly aged >80 and with stroke had a higher (although not significant) probability of falling if randomized in the IG (RR=1.21, 95%CI [0.65, 1.95] and, respectively, RR=1.21, 95%CI [0.51, 2.83]).

Considering the endpoint ‘time to the first fall’, elderly aged 65-80, elderly aged >80, and elderly with Parkinson’s had a lower (although not significant) hazard ratio if randomized in the IG in comparison to the CG (HR= 0.83, 95%CI [0.56, 1.25]); HR=0.92, 95%CI [0.54, 1.59]; HR=0.83, 95%CI [0.49, 1.41], respectively), Instead, the hazard ratio was higher (HR=1.32, 95%CI [0.58, 3.01]) for elderly with stroke sequelae randomized in the IG.

### 3.4 Rasch analysis

The final solutions for both the FROP-Com and the Balance scales showed adequate fitness to the Rasch Model (Table 6). Hence, it was possible to devise conversion tables from ordinal scores to interval-level measurements (having, as a unit of measurement, the logit), then used for the subsequent analysis.

**Table 6.**
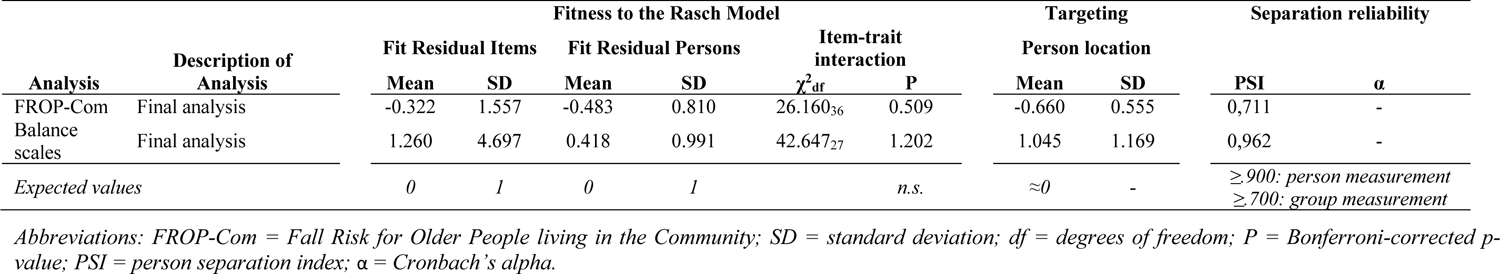
Rasch analysis results (final analyses) on scales used for pre-test vs. post-test differences analysis

| Analysis | Description of Analysis | Fitness to the Rasch Model |  |  |  |  |  | Targeting |  | Separation reliability |  |
| --- | --- | --- | --- | --- | --- | --- | --- | --- | --- | --- | --- |
| | | Fit Residual Items | | Fit Residual Persons | | Item-trait interaction | | Person location | | PSI | $\alpha$ |
| | | Mean | SD | Mean | SD | $\chi^2_{df}$ | P | Mean | SD | | |
| FROP-Com | Final analysis | -0.322 | 1.557 | -0.483 | 0.810 | 26.160<sub>36</sub> | 0.509 | -0.660 | 0.555 | 0,711 | - |
| Balance scales | Final analysis | 1.260 | 4.697 | 0.418 | 0.991 | 42.647<sub>27</sub> | 1.202 | 1.045 | 1.169 | 0,962 | - |
| Expected values |  | 0 | 1 | 0 | 1 | n.s. |  | ≈0 | - | ≥ 900: person measurement<br>≥ 700: group measurement |  |

Abbreviations: FROP-Com = Fall Risk for Older People living in the Community; SD = standard deviation; df = degrees of freedom; P = Bonferroni-corrected p- value; PSI = person separation index; $\alpha$ = Cronbach's alpha.

### 3.5 Pre-test vs. post-test differences between groups (CG and IG)

The ANCOVA analysis (table 7) showed no significant difference between the CG and the IG for the post-test FROP-Com (MD=-0.03 logits, 95%CI [-0.13, 0.07]), BBS (+0.15 logits, 95%CI [-0.13, 0.07]), and POMA measures (+0.12 logits, 95%CI [-0.14, 0.37]) after controlling for the pre-test values.

**Table 7.** Analysis of pre-test vs. post-test differences for FROP-Com and balance indicators

| Variable |  | Control group |  |  | Intervention group |  |  | Comparison |  | Effect size<br>Cohen's d (CI95%) |
| --- | --- | --- | --- | --- | --- | --- | --- | --- | --- | --- |
|  |  | mean | sd | n | mean | sd | n | MD (CI95%) | p |  |
| FROP-Com | pre | -0.5 | 0.5 | 200 | -0.5 | 0.5 | 203 |  |  |  |
|  | post | -0.9 | 0.6 | 132 | -0.9 | 0.6 | 153 |  |  |  |
|  | post - pre | -0.3 | 0.4 | 132 | -0.3 | 0.4 | 153 | -0.03 (-0.13; 0.07) | 0.543 | -0.08 (-0.32; 0.15) |
| BBS | pre | 2.2 | 1.6 | 200 | 2.1 | 1.6 | 203 |  |  |  |
|  | post | 2.5 | 1.8 | 156 | 2.6 | 1.8 | 184 |  |  |  |
|  | post - pre | 0.1 | 2.7 | 156 | 0.5 | 2.3 | 184 | 0.15 (-0.23; 0.53) | 0.445 | 0.14 (-0.08; 0.35) |
| POMA | pre | 2.6 | 1.8 | 200 | 2.6 | 1.9 | 203 |  |  |  |
|  | post | 3.0 | 1.9 | 156 | 3.1 | 1.9 | 182 |  |  |  |
|  | post - pre | 0.2 | 1.2 | 156 | 0.3 | 1.3 | 182 | 0.12 (-0.14; 0.37) | 0.363 | 0.10 (-0.11; 0.31) |
| MBT | pre | 0.6 | 2.3 | 198 | 0.4 | 2.4 | 200 |  |  |  |
|  | post | 1.0 | 2.5 | 156 | 1.2 | 2.7 | 182 |  |  |  |
|  | post - pre | 0.2 | 1.8 | 155 | 0.6 | 2.0 | 179 | **0.42 (0.03; 0.81)** | **0.035** | **0.26 (0.04; 0.48)** |
| FABS | pre | 0.4 | 1.2 | 200 | 0.4 | 1.2 | 203 |  |  |  |
|  | post | 0.4 | 1.2 | 156 | 0.4 | 1.3 | 184 |  |  |  |
|  | post - pre | 0.1 | 0.7 | 156 | 0.3 | 0.7 | 184 | **0.21 (0.06; 0.36)** | **0.006** | **0.32 (0.10; 0.53)** |

Abbreviations: sd = standard deviation; MD = mean differences in post-test values adjusted for pre-test values; CI95% = Confidence Interval at 95% level; FROP- Com = Fall Risk for Older People in the Community; BBS = Berg Balance Scale; POMA = Performance Oriented Mobility Assessment; FABS = Fullerton Advanced Balance Scale; MBT = Mini-BESTest.
Notes: in bold, significant pre-test vs. post-test differences were reported.

However, there were statistically significant differences for the post-test MBT measures (MD=+0.42 logits, 95%CI [0.03, 0.81], p=0.035; effect size 0.26, 95%CI [0.04; 0.48]) and FABS (MD=+0.21 logits, 95%CI [0.06, 0.36], p=.006; effect size 0.32, 95%CI [0.10; 0.53]) between the two groups after controlling for the pre-test measures.

## 4 Discussion

In the PRE.C.I.S.A. RCT study, we evaluated the efficacy of the simultaneous administration of a multiple-component and a multifactorial personalized intervention in reducing fall rates in community-dwelling older adults at moderate-to-high fall risk compared to usual care. Another innovative aspect of the study was the inclusion in the sample of elderly with an even higher fall risk because of a concomitant diagnosis of Parkinson’s Disease or stroke sequelae. The results showed no statistically significant differences between groups regarding the number of falls, the falling probability, and the time to the first fall at a twelve-month follow-up. According to the subgroup analysis, no significant differences were reported between groups. However, a lower number of falls, lower fall rates in multiple fallers, a lower mean number of falls per participant, and a lower rate of fall-related severe injuries were recorded for the intervention group, although the differences were not significant. Finally, a significant improvement for two balance-related indicators at post-test was recorded in the intervention after controlling for the pre-test measures.

Several previous studies investigated the effects of different interventions on fall prevention in community-dwelling older adults. However, their effectiveness in reducing falls and their highly disabling consequences is still controversial (37–39). Moreover, no evidence was available on simultaneously multiple-component and multifactorial interventions to manage fall risk (12). Furthermore, RCTs aiming at reducing falls in the elderly usually exclude those with an even higher risk of falling because of an associated neurological condition, such as Parkinson’s Disease (PD) and stroke. Thus, this trial was built upon two innovative hypotheses. First, we hypothesized that combining a multiple-component intervention with a personalized multifactorial intervention could reduce fall rates in community-dwelling older adults. Secondly, considering that most of the risk factors for falling are independent of the diseases associated with falls, we hypothesized that most of those risk factors could be targeted by the same interventions independently from the participant’s risk class. Considering that the devised multicomponent intervention also included elements of physical exercise that were disease-specific, we were able to enroll the elderly with neurological conditions who were at a high risk of falling, such as those affected by PD and stroke sequelae (40–48,48)

According to our results, the total number of recorded falls was substantial (690), with a fall incidence of 58.6% and a mean of 1.71 (SD: 3.36) falls accounted for each included subject. These data are in contrast with those reported in the literature, where the overall fall incidence in elderly over 65 years is around 28 to 35% and about 32 to 42% in those over 75 years (49), with 0.2-1.6 falls for each included subject (2). Furthermore, the prevalence of ‘multiple fallers’ observed in our trial (22.1%) was higher than that reported by previous studies (15%) in older adults (45). Indeed, several subjects reported more than ten falls in our sample while participating in the trial, with a subject reporting up to thirty-three falls (2.8 falls per month). The observed discrepancies between our results and literature data may be explained considering the inclusion of participants affected by neurological conditions who are well known to be multiple fallers and present, according to previous studies, a fall risk of around 50% in Parkinson’s Disease (50) and 43 to 70% in stroke (51). Indeed, the literature’s reported incidence of multiple fallers among persons with neurological conditions is around 15% in stroke subjects (51) and over 50% incidence in PD, where up to 13% of patients fall more than once a week (45). The prevalence of severe injuries in our sample (6.8% in the CG and 5.9% in the IG; 6.4% for the whole sample) was instead similar to the value (10%) reported for the elderly population (49).

The statistical analyses revealed no significant differences in fall rates and related parameters between IG and CG, i.e., fall severity, fall probability for one, two or more, and three or more falls within twelve months, and time to the first fall. These results align with Lamb’s and Cattaneo’s works (13, 37). However, a systematic (although not significant) trend of better outcomes was reported for the IG compared to the CG. In particular, we recorded fewer falls, lower fall rates in multiple fallers, a lower mean number of falls per participant, and a lower rate of fall-related severe injuries in the IG. Furthermore, considering the falling probability for three or more falls (multiple fallers), it was in favor of the IG group slightly above the chosen level of statistical significance. The subgroup analysis yielded similar results, with the absence of significant differences between the CG and IG regarding the number of falls, the falling probability, or the time to the first fall (all p>0.05) across all the four considered subgroups (older adults 65 to 79 years; older adults ≥80 years; older adults with PD; older adults with stroke). As per the general group, there was a general, although not significant, trend of better outcomes for the IG group. In particular, if we consider the falling probability for two or more falls in the elderly with PD, this was very close to the level of statistical significance. The stroke subgroup made an exception, as the risk of falling was apparently higher (although not significant) for patients enrolled in the IG. The fall risk for persons with stroke is notoriously high, being reported to be around 43 to 70% in the previous trials (51). Undoubtedly, intravenous thrombolysis (52) and endovascular treatments for large vessel occlusions (53) have significantly improved the survival and long-term functional outcome of ischemic stroke. However, their impact on balance and other fall risk factors is unclear. Our results could also be explained in light of the literature data, which report an increase in the exposure to circumstances leading to falls (and, thus, an increase in the number of falls) brought about by an increase in physical activity (54).

The analysis of the differences between baseline (T1) and 3-month follow-up (T3) across CG and IG was preceded by a Rasch Analysis. The latter was performed because scales’ total scores are ordinal in nature and, as such, should not be used with parametric statistical techniques such as ANCOVA (55, 56), as it may lead to erroneous results (57). The Rasch analysis allowed to elaborate conversion tables of the scales’ total scores into invariant interval-level estimates of ability (whose unit of measurement is the logit) that satisfy the mathematical requirements of a general measurement theory called Additive Conjoint Measurement (58, 59). In other words, interval-level estimates produced by Rasch analysis are comparable in measurement properties to those delivered by instruments measuring physical variables, such as a thermometer. Thus, those interval-level estimates were employed for the subsequent analysis of covariance. After adjusting for the baseline values, the post-test measurements of a comprehensive fall risk indicator (FROP-Com) and four balance scales were compared between the CG and IG. In this way, we were able to compare the differences between groups ascribable to the administered intervention without introducing any bias due to the use of ordinal metrics with a parametric statistical method such as ANCOVA, which requires continuous measures.

In a previous study evaluating the effects of a home-based exercise program in reducing falls in the elderly population, Vogler et al. observed an improvement in reducing fall risk and balance indicators at the end of the twelve-week treatment and a subsequent return to baseline values after 24 weeks (60). Indeed, the ANCOVA showed a significant effect on balance within the intervention group only for two of the four balance indicators (FABS and Mini-BESTest). This result could be explained considering that the latter indicators are more challenging in balance ability than BBS and POMA (30, 61). In other words, no effect was likely shown with BBS and POMA because the ability range of the sample was higher than the difficulty level of the two scales. Despite the short-term effectiveness of the intervention, the small effect size of the balance change suggests that the ratio between treatment benefits and costs of administering physical exercise for the overall study duration (12 months), as suggested by some authors (60), may be unfavorable.

Indeed, the main hypothesis behind this study was that all the proposed interventions could contribute equally to avoiding the detraining effect by facilitating the adoption of a habit of regularly performing exercise and physical activity. However, the study results seem to contradict this hypothesis. First, there was no significant reduction of the overall burden of fall risk factors on post-test, as shown by the results of ANCOVA performed on FROP-Com. Second, participants in the IG provided informal positive feedback on some but not all the interventions. Indeed, the activities involving social participation, such as group exercise, educational sessions, and physiotherapy home visits, were particularly appreciated. This could be explained considering that these activities also offered socializing opportunities, thus contrasting the social isolation, which, per se, might be a fall risk factor (62) and may have a highly significant negative impact on the health and wellbeing of older people (63). At the same time, this could be an indirect indication of a lower appreciation of the home physical exercise program. Finally, if the home exercise program was not integrated early into the participants’ daily routine, this could have led, in turn, to a lower engagement at home. The latter may have facilitated a detraining effect after the eleven treatment weeks, thus losing any eventual long-term beneficial effect of the combined treatment strategy.

### 4.1 Study limitations

The present study results should be considered in light of some limitations. First, although the physical activity report diaries suggested a lower adherence to physical exercise in the home setting for the Intervention Group, it cannot be excluded that the information provided by participants was not reliable enough. For this reason, the use of wearable sensors (64), i.e., portable inertial measurement units, to continuously record the subject’s activity and adherence to home physical exercise, together with remote telemedicine support, could be strategic to achieving participants’ compliance to treatment and better monitor executed exercises. Further studies should be conducted to explore fall risk management properly in a similar scenario.

Second, the home fall diaries reporting was not reliable enough. Participants were expected to report each occurred fall accurately in the diary, but this might have happened only partially. As previously experienced in other trials, many participants reported falls inconsistently. This difficulty of older subjects in recalling is well documented in the literature, turning out to be underestimating single falls and overestimating multiple falls (9, 65). To prevent this bias, completing a daily diary was tested (66); however, people reporting a high number of falls turned out not to return the diary at the end of the trial (67). Different options were proposed in the literature to deal with this issue: monthly diary return through postal service (66), monthly follow-up calls to punctually record falls (68), incentives for monthly diary return (69), and personalization of the latter (69). In this study, a monthly follow-up call was performed to investigate falls recorded in the previous month, but these data did not match those observed at the twelve-month follow-up when diaries were returned. Moreover, several participants did not return the diary or filled it in only partially. Thus, data recorded during monthly phone calls were used to estimate the fall rate for statistical analysis.

Finally, we observed limited participation with a higher drop-out rate and low adherence to trial post-test assessment in the Control Group. This might have biased the ANCOVA results at the post-test. Therefore, future trials should consider offering a placebo treatment to avoid an excessive loss to follow-up in the control group.

### 4.2 Conclusions

This trial attempted to provide a new concept of intervention to reduce falls in a mixed population of older people. The intervention showed the potentiality of improving balance at post-test, leading to a positive trend toward a lower number of falls, lower fall rates in multiple fallers, a lower mean number of falls per participant, and a lower rate of fall-related severe injuries for the intervention group. However, as these differences were not significant, the proposed intervention must be considered ineffective in reducing the number of falls, the falling probability, and the time to the first fall at the twelve-month follow-up in community-dwelling older adults at moderate-to-high fall risk.

Unfortunately, other recent RCTs have reached similar conclusions for other interventions (13,70,71). Therefore, the temptation would be high to sustain that, as there are no effective interventions, no further efforts should be made to prevent falls and fall-related injuries and improve safe physical mobility in our aging societies. Indeed, as proposed by a recent commentary (72), there is a need for new, better concepts to increase the efficacy of interventions to reduce falls and their consequences. In this respect, the widespread use of ICT solutions could represent an opportunity to be explored. For instance, regarding this trial, the results might have been different if an ICT-based solution could be adopted to monitor the participants’ activity levels and record any eventual falls that occurred automatically. In this way, we would have overcome the limitations imposed by the unreliability of the fall diaries.

We believe that future studies exploring different effects of combined multiple-component and personalized multifactorial interventions to reduce falls and subsequent consequences should be planned with a clear plan for overcoming the limitations highlighted in the PRE.C.I.S.A. study.

## Authors’ contributions

All authors contributed equally to the manuscript and read and approved the final version of the manuscript.

## Data Availability

All data produced in the present study are available upon reasonable request to the authors

## Acknowledgments

The authors are grateful to all professionals who collected data during the PRE.C.I.S.A. study.

## PRECISA study group members

### Ospedale Civile di Baggiovara (Modena, Italy)

- *Rehabilitation Medicine:* Fabio La Porta, Serena Caselli, Pierina Viviana Clerici, Stefano Cavazza, Valeria Serraglio, Maria Cristina Vannini, Federica Bovolenta, Giada Lullini, Silvia Puglisi, Angela Gallo
- *Elderly Medicine:* Chiara Mussi, Marco Bertolotti, Roberto Scotto, Giulia Lancellotti
- *Neurology:* Franco Valzania, Francesca Falzone
- *Primary Care Department:* Monica Montanari, Maria Luisa De Luca, Emanuela Malagoli, Elisa Franchini
- *Research and Innovation Unit:* Luisa Palmisano, Franca Serafini

### Arcispedale Santa Maria Nuova (Reggio Emilia, Italy)

- *Rehabilitation Medicine:* Claudio Tedeschi, Gioacchino Anselmi, Valentina D’Alleva, Mariangela Di Matteo, Rosalinda Ferrari, Stefania Costi, Filomena Simeone, Giulia D’Apote, Alessandra Rizzica, Maria Beatrice Galavotti, Marta Ghirelli
- *Elderly Medicine:* Giulio Pioli, Chiara Bendini, Giulia Lancellotti
- *Neurology:* Massimo Bondavalli, Eleni Georgopoulos

### University of Modena and Reggio Emilia, Unit of statistical and methodological support for clinical research

- Roberto D’Amico, Sara Balduzzi, Roberto Vicini, Federico Banchelli

### University of Bologna, Department of Electrical, Electronic, and Information Engineering ‘Guglielmo Marconi’ (DEI)

- Lorenzo Chiari, Sabato Mellone, Alice Coni

## Supplementary materials

The Supplementary Material 1 and 2 for this article can be found online at …

### Contribution to the field statement

The PRE.C.I.S.A. trial attempted to provide a new concept of intervention to reduce falls in a mixed population of older people, including the elderly with an even higher fall risk because of a concomitant diagnosis of Parkinson’s Disease or stroke sequelae. The intervention showed the potentiality of improving their balance, leading to a positive trend toward a lower number of falls, lower fall rates in multiple fallers, a lower mean number of falls per participant, and a lower rate of fall-related severe injuries for the intervention group. However, as these differences were not statistically significant, the proposed intervention must be considered ineffective in reducing the fall rates at the twelve-month follow-up in a mixed population of community-dwelling older adults at moderate-to-high fall risk. As similar RCTs have recently reached similar conclusions for other interventions, there is a need for new, better concepts to increase the efficacy of interventions to reduce falls and their consequences. In this respect, the widespread use of ICT solutions could represent an excellent opportunity to be explored.

## Supplementary Material 1 Group intervention protocol details

As described in the Methods section, participants in the IG were taken in charge by an interdisciplinary team including a Physiatrist (P), a Physiotherapist (PT), a Geriatrician (G), and a Neurologist (N) who administered synergically the following five interventions.

**1. Group exercise sessions:** one weekly six-person group session of sixty minutes for eleven weeks. Each session was composed of the following parts:
  i. Warming-up (five minutes): head, neck, trunk, and ankle movements, back and knee extensions, walking on the spot;
  ii. Circuit training (thirty-five minutes):
    - Station 1 - Muscular strength exercises: ankle plantarflexion, squat, chair standing, and frontal step (all these exercises were realized with hand support and, when possible, wearing a weight vest).
    - Station 2 - Balance exercises: reaching in standing position, tandem standing, and single-leg standing (1-6 weeks); sidewalk, tandem walk, and toe walk (7 - 11 weeks).
    - Station 3 - Recovery techniques from falling: seven steps with the ‘backward-chaining method’, starting from the sitting or standing position(1).
  i. Dynamic balance and walking, considering the base pathology (ten minutes):
    - Obstacle courses (walking with motor and dual cognitive tasks);
    - Walking exercises (direction and speed changes, associated activities with arms, in crowded contexts; in PD use of visual and auditory cues);
    - Climbing stairs.

The remaining ten minutes were destined to rest and control the physical activity report diary. During the first session, the Physiotherapist delivered a weight vest to each IG participant and verified the initial level for each of the cited three circuit training stations. Specifically:

- Station 1 - Muscular strength exercises:
  - First session: for each exercise and each side (in the case of bilateral exercises), the Physiotherapist calculated the subject’s maximum weight (1RM) based on the highest number of consecutive repetitions performed by him/her (starting from 1,5 kg, then calculate the 1RM and the percentage of 65% of the 1RM using ad hoc tables built on the average result of three spread formulas (Brzycki, Baechle, Epley)) and noted it in the participant’s group treatment chart.
  - Third-Fifth-Seventh-Ninth session: for each exercise and side, the Physiotherapist checked the possible progression (starting from the current working weight, increasing progressively 500 grams to recalculate the 65% of 1RM as described) and noted it in the participant’s group treatment chart.

During the 1RM, the participant had to work to a CR-10 Borg Scale level of ‘10 – extremely strong’, while during muscular strength group exercise with a CR-10 Borg Scale level of ‘5 – Strong’.

- Station 2 – Balance exercises:
  - First session: for each exercise, the Physiotherapist defined the initial difficulty level (4 levels A-B-C-D of increasing difficulty). He started from medium-high C level, then receded or advanced according to the subject’s condition verified during pre-test evaluations.
  - Third-Fifth-Seventh-Ninth session: for each exercise, the Physiotherapist checked the possible progression to the next level, which could occur if the participant realized the activity in a safe condition.

- Station 3 – Recovery techniques from falling:
  - In the first session, the Physiotherapist proposed performing steps 1 and 2, then further 5 steps (to 7), one for each session.
  - It was recommended that the Physiotherapist review the steps of the previous weeks and set the next step for every session.
  - The subject passed to the next step only when he/she could safely perform the previous steps.

At each group session, the participant must have his/her weight vest, his/her fall-physical activity report diary of the current month, and his/her manual of the home exercise program (see next point 4).

During rest periods from exercises, the participant delivered his/her fall-physical activity report diary and, in case of at least one fall during the week, the completed ‘fall report’. In addition, at the end of the session, the Physiotherapist updated the manual of the home exercise program with the weekly level progression of the exercises (the passage changed every two weeks, but depending on individual needs, it was possible to add other series of the same exercise in the intermediate weeks).

**2. Group education sessions on fall risk factors:** one weekly thirty-minute session (held after the group exercise session) for eleven weeks focused on modifiable fall risk factors and risky behaviors. Two parts constituted each session: a ten-minute frontal lesson on a specific theme held by a component of the interdisciplinary team, followed by a twenty-minute group discussion on the lesson content (participants, their caregivers, and the professional). During the eleven weeks, the following topics were proposed:
  i. Why is it important to prevent falls? Why do you fall? (G)
  ii. The importance of regular physical exercise (P)
  iii. Behaviors at risk of falling (PT)
  iv. Drugs and falls (G)
  v. Home safety (PT)
  vi. Postural hygiene of feet and footwear (PT)
  vii. The benefits of a healthy and proper diet and the adoption of healthy lifestyles in fall prevention (G)
  viii. Attention and falls (N)
  ix. Vision impairment and falls (G)
  x. Osteoporosis and falls (G)
  xi. Why is it important to continue exercising? (PT)

A handbook summarizing these topics was provided to each participant at the beginning of the first education session.

**3. A personalized plan of reducing domestic fall risk factors following a home visit performed by the Physiotherapist within the first week of treatment.** During this visit, lasting sixty-ninety minutes, the Physiotherapist filled out the ‘Home environmental risks questionnaire’ and compared it with the same questionnaire compiled by the participant at the pre-test assessment. This questionnaire investigated the following macro-areas related to modifiable risk factors:
  - Slippery floors (outside the house, inside the house);
  - Stairs (handrail, height, tread, scale width);
  - Lighting;
  - Carpets, doormats, movable floor coverings, electric wires;
  - Isolated steps, important disconnections;
  - Furniture and ornaments (chairs, table, protruding feet);
  - Objects at high height;
  - Bed height;
  - Bathroom fixtures (toilet height, shower/bath surfaces).

Then, the Physiotherapist gave specific recommendations with proposals for correcting the detected modifiable risk factors by delivering the ‘Suggestions for the reduction of environmental risks at home’ sheet in which the actual present hazards were highlighted. He/she also verified the presence of the fall-physical activity report diary, delivered at the time of recruitment, in a position that facilitated the compilation in case of fall (e.g., hanging on the wall in the living room/ kitchen, etc.).

Finally, during the following three home visits related to the personalized home exercise program, the Physiotherapist checked the implementation of recommended corrections in the first home access and filled the ‘Check-list of correction of environmental risk factors at home’.

**4. A personalized home exercise program**, coordinated with the group exercise program aimed at improving strength, static and dynamic balance, and acquiring a long-term daily habit of exercise and physical activity in the context of a progressive and permanent adoption of a healthy and active lifestyle. The Physiotherapist devised this program in the context of an initial home visit (2^nd^ week) and subsequently monitored within two further home visits (4^th^ and 6^th^ weeks). During the initial home visit, an illustrated manual containing strength and balance exercises was provided and explained to each participant, based on the first group exercise session (in which the subject’s maximum weight (1RM) for strength exercises and the initial difficulty level for balance exercises were calculated). These exercises were chosen between those the subject performed with greater safety in the group session. The indication given to the subject was to:
  - Perform muscular strength and balance exercises two more times a week, in addition to the group session performed at the center, for about thirty minutes, possibly on alternate days to allow rest between one session and the following;
  - Perform a thirty-minute walking session on rest days from exercises, at least twice a week. These sessions could be divided into shorter sessions (e.g., at the beginning three ten-minute sessions, alternate with rest, and then increase reaching a single thirty-minute session);
  - Register the performed physical activity in his/her fall-physical activity report diary of the current month, which the Physiotherapist checked during the weekly group session.

During the subsequent two visits, the Physiotherapist verified the setting adequacy and modality in which the participant performed the suggested exercises and updated the fall-physical activity report diary.

Finally, in all the three home access linked to the home exercise program, the Physiotherapist checked the implementation/maintenance of the recommendations on risk factors correction given in the first-week home visit.

**5. A multifactorial personalized intervention aiming at modifying additional risk factors**
which were performed by the interdisciplinary team and included the following interventions:
  - Review of medications, including psychotropic medications (N and G), antiparkinsonian drugs (N), and cardiovascular medications (G);
  - Management of unaddressed visual impairments (G): ophthalmologist referral, lens prescription, suggestions regarding the limitation of bifocal lenses;
  - Management of unaddressed cardiovascular issues (G), such as postural hypotension, covert cardiac failure, and abnormalities of cardiac rhythm, eventual cardiology referral;
  - Vitamin D prescription (G);
  - Improvement of nutritional state (G), with prescription of caloric-proteic integration and/or nutritional referral;
  - Management of muscle-skeletal issues, including spasticity (P and PT);
  - Education about foot self-care, including podologist referral if appropriate (P);
  - Assessment, prescription, and final testing of orthesis and mobility aids, including proper shoes (P and PT).

Interventions one to four were administered to all IG participants (multiple-component intervention), whereas the multifactorial intervention (intervention five) was personalized based on the individual fall risk profile devised on the pre-test assessment. Furthermore, interventions one, two, and five were conducted within an outpatient setting, while interventions three and four were home-based.

## Supplementary Material 2 Outcome measure details (AAE and OAE assessments)

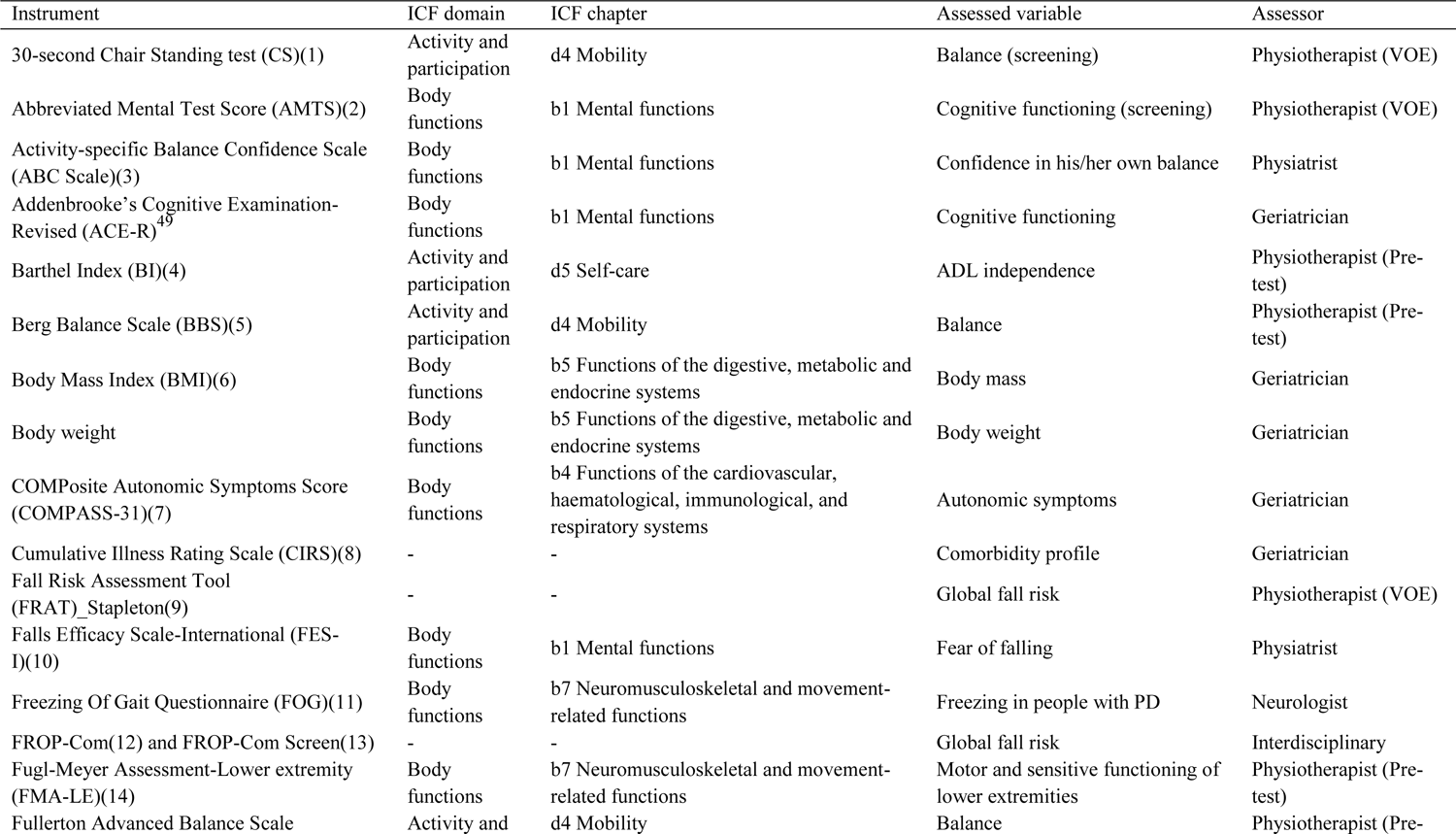
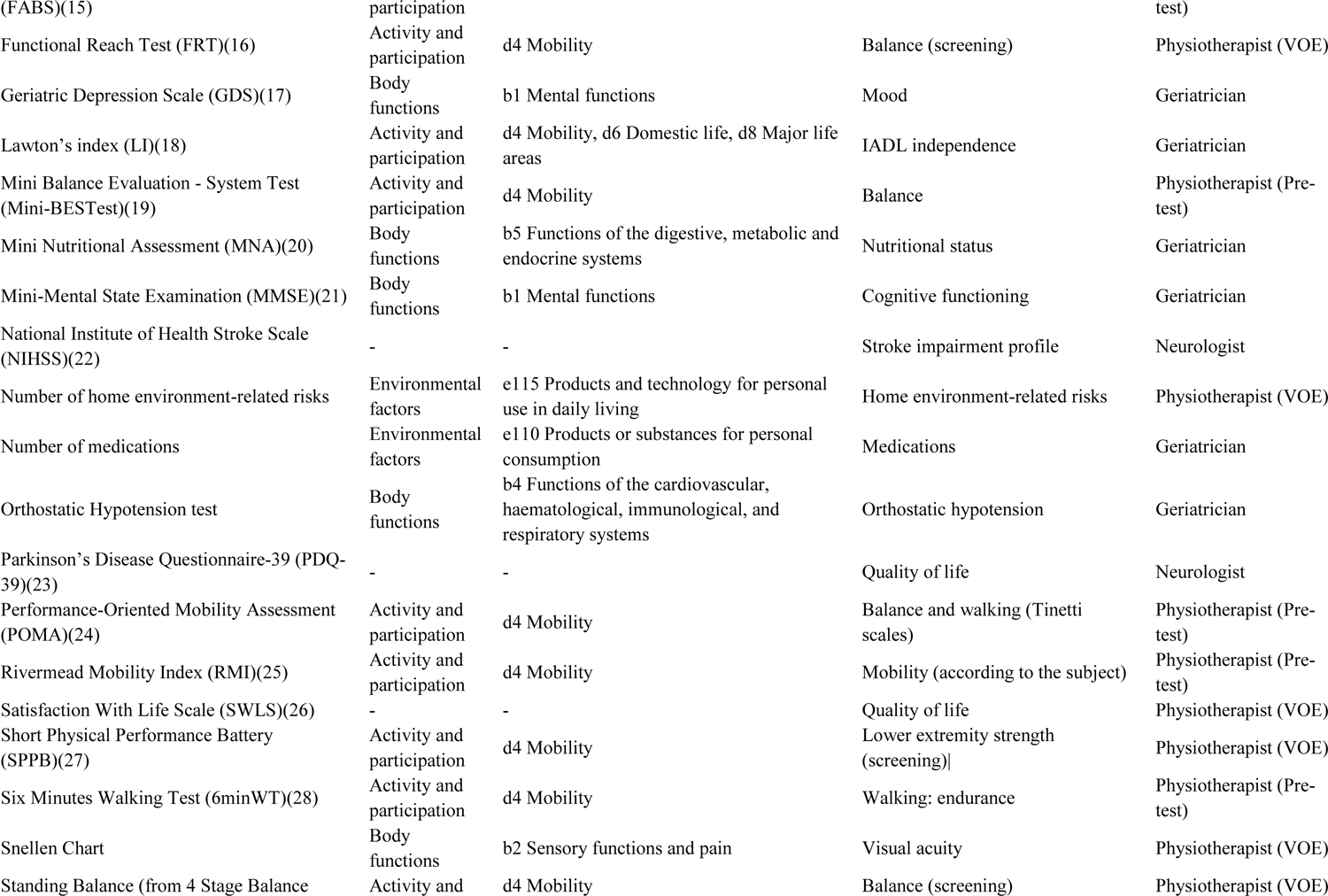
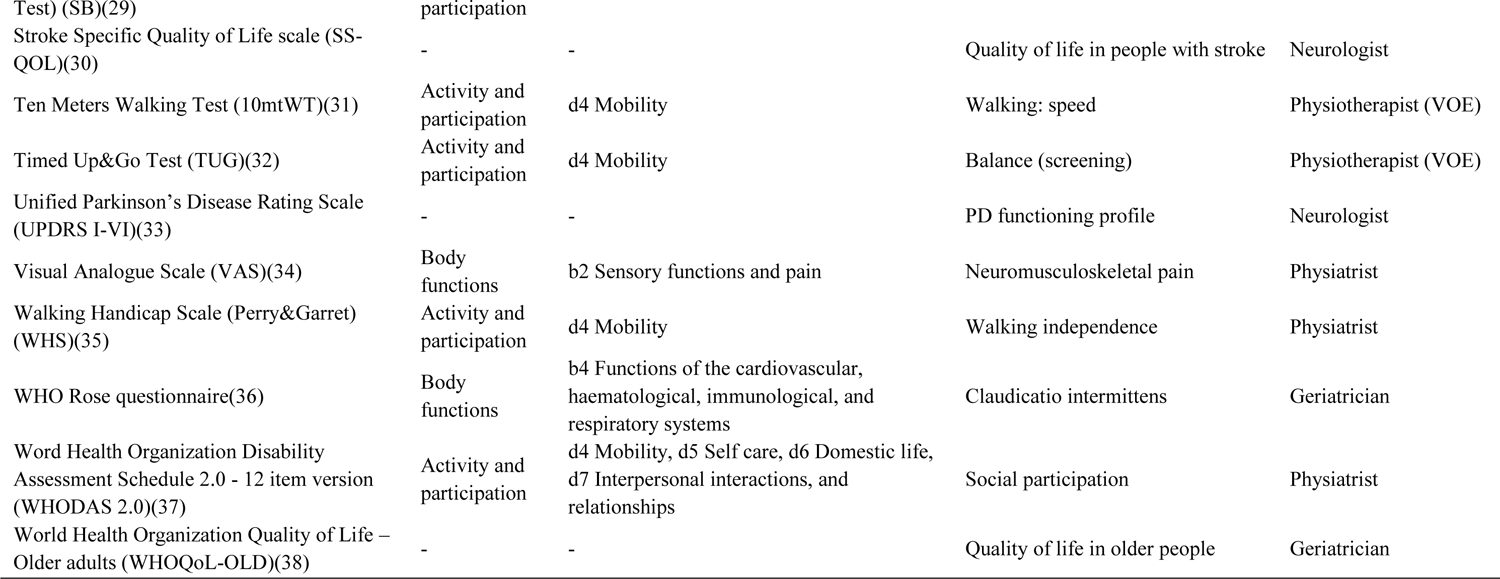

*Supplementary Material 2 - Outcome measure details (AAE and OAE assessments)*
| Instrument | ICF domain | ICF chapter | Assessed variable | Assessor |
| --- | --- | --- | --- | --- |
| 30-second Chair Standing test (CS)(1) | Activity and participation | d4 Mobility | Balance (screening) | Physiotherapist (VOE) |
| Abbreviated Mental Test Score (AMTS)(2) | Body functions | b1 Mental functions | Cognitive functioning (screening) | Physiotherapist (VOE) |
| Activity-specific Balance Confidence Scale (ABC Scale)(3) | Body functions | b1 Mental functions | Confidence in his/her own balance | Physiatrist |
| Addenbrooke's Cognitive Examination-Revised (ACE-R)<sup>49</sup> | Body functions | b1 Mental functions | Cognitive functioning | Geriatrician |
| Barthel Index (BI)(4) | Activity and participation | d5 Self-care | ADL independence | Physiotherapist (Pre-test) |
| Berg Balance Scale (BBS)(5) | Activity and participation | d4 Mobility | Balance | Physiotherapist (Pre-test) |
| Body Mass Index (BMI)(6) | Body functions | b5 Functions of the digestive, metabolic and endocrine systems | Body mass | Geriatrician |
| Body weight | Body functions | b5 Functions of the digestive, metabolic and endocrine systems | Body weight | Geriatrician |
| COMPOSITE Autonomic Symptoms Score (COMPASS-31)(7) | Body functions | b4 Functions of the cardiovascular, haematological, immunological, and respiratory systems | Autonomic symptoms | Geriatrician |
| Cumulative Illness Rating Scale (CIRS)(8) | - | - | Comorbidity profile | Geriatrician |
| Fall Risk Assessment Tool (FRAT)_Stapleton(9) | - | - | Global fall risk | Physiotherapist (VOE) |
| Falls Efficacy Scale-International (FES-I)(10) | Body functions | b1 Mental functions | Fear of falling | Physiatrist |
| Freezing Of Gait Questionnaire (FOG)(11) | Body functions | b7 Neuromusculoskeletal and movement-related functions | Freezing in people with PD | Neurologist |
| FROP-Com(12) and FROP-Com Screen(13) | - | - | Global fall risk | Interdisciplinary |
| Fugl-Meyer Assessment-Lower extremity (FMA-LE)(14) | Body functions | b7 Neuromusculoskeletal and movement-related functions | Motor and sensitive functioning of lower extremities | Physiotherapist (Pre-test) |
| Fullerton Advanced Balance Scale | Activity and | d4 Mobility | Balance | Physiotherapist (Pre- |
| (FABS)(15) | participation |  |  | test) |
| Functional Reach Test (FRT)(16) | Activity and participation | d4 Mobility | Balance (screening) | Physiotherapist (VOE) |
| Geriatric Depression Scale (GDS)(17) | Body functions | b1 Mental functions | Mood | Geriatrician |
| Lawton's index (LI)(18) | Activity and participation | d4 Mobility, d6 Domestic life, d8 Major life areas | IADL independence | Geriatrician |
| Mini Balance Evaluation - System Test (Mini-BESTest)(19) | Activity and participation | d4 Mobility | Balance | Physiotherapist (Pre-test) |
| Mini Nutritional Assessment (MNA)(20) | Body functions | b5 Functions of the digestive, metabolic and endocrine systems | Nutritional status | Geriatrician |
| Mini-Mental State Examination (MMSE)(21) | Body functions | b1 Mental functions | Cognitive functioning | Geriatrician |
| National Institute of Health Stroke Scale (NIHSS)(22) | - | - | Stroke impairment profile | Neurologist |
| Number of home environment-related risks | Environmental factors | e115 Products and technology for personal use in daily living | Home environment-related risks | Physiotherapist (VOE) |
| Number of medications | Environmental factors | e110 Products or substances for personal consumption | Medications | Geriatrician |
| Orthostatic Hypotension test | Body functions | b4 Functions of the cardiovascular, haematological, immunological, and respiratory systems | Orthostatic hypotension | Geriatrician |
| Parkinson's Disease Questionnaire-39 (PDQ-39)(23) | - | - | Quality of life | Neurologist |
| Performance-Oriented Mobility Assessment (POMA)(24) | Activity and participation | d4 Mobility | Balance and walking (Tinetti scales) | Physiotherapist (Pre-test) |
| Rivermead Mobility Index (RMI)(25) | Activity and participation | d4 Mobility | Mobility (according to the subject) | Physiotherapist (Pre-test) |
| Satisfaction With Life Scale (SWLS)(26) | - | - | Quality of life | Physiotherapist (VOE) |
| Short Physical Performance Battery (SPPB)(27) | Activity and participation | d4 Mobility | Lower extremity strength (screening) | Physiotherapist (VOE) |
| Six Minutes Walking Test (6minWT)(28) | Activity and participation | d4 Mobility | Walking: endurance | Physiotherapist (Pre-test) |
| Snellen Chart | Body functions | b2 Sensory functions and pain | Visual acuity | Physiotherapist (VOE) |
| Standing Balance (from 4 Stage Balance | Activity and | d4 Mobility | Balance (screening) | Physiotherapist (VOE) |
| Test) (SB)(29) | participation |  |  |  |
| Stroke Specific Quality of Life scale (SS-QOL)(30) | - | - | Quality of life in people with stroke | Neurologist |
| Ten Meters Walking Test (10mtWT)(31) | Activity and participation | d4 Mobility | Walking: speed | Physiotherapist (VOE) |
| Timed Up&Go Test (TUG)(32) | Activity and participation | d4 Mobility | Balance (screening) | Physiotherapist (VOE) |
| Unified Parkinson's Disease Rating Scale (UPDRS I-VI)(33) | - | - | PD functioning profile | Neurologist |
| Visual Analogue Scale (VAS)(34) | Body functions | b2 Sensory functions and pain | Neuromusculoskeletal pain | Physiatrist |
| Walking Handicap Scale (Perry&Garret) (WHS)(35) | Activity and participation | d4 Mobility | Walking independence | Physiatrist |
| WHO Rose questionnaire(36) | Body functions | b4 Functions of the cardiovascular, haematological, immunological, and respiratory systems | Claudicatio intermittens | Geriatrician |
| Word Health Organization Disability Assessment Schedule 2.0 - 12 item version (WHODAS 2.0)(37) | Activity and participation | d4 Mobility, d5 Self care, d6 Domestic life, d7 Interpersonal interactions, and relationships | Social participation | Physiatrist |
| World Health Organization Quality of Life – Older adults (WHOQoL-OLD)(38) | - | - | Quality of life in older people | Geriatrician |

